## Supplementary material for "Estimating lengths-of-stay of hospitalized COVID-19 patients using a non-parametric model: a case study in Galicia (Spain)"

**ORCIDs**: Ana López-Cheda (<https://orcid.org/0000-0002-3618-3246>)^1^, María-Amalia Jácome (<https://orcid.org/0000-0001-7000-9623>)^1^, Ricardo Cao ([https://orcid.org/0000-0001-8304-687X)^2^](https://orcid.org/0000-0001-8304-687X)2), Pablo M De Salazar ([https://orcid.org/](https://orcid.org/0000-0001-8304-687X)2)0000-0002-8096-2001)^3^

**This Supplementary Material includes:**

Appendix text

Figures and Tables

Script (R) for reproducing the results

**S1. Study population and sample description**

The study population comprised a total of 10454 COVID-19 reported cases in Galicia (Spain) between March and May 2020. From these subjects, only 2484 were admitted to the hospital, though 31 of them were discharged or died on the same day. This study includes only the 2453 patients admitted to the hospital for at least one day. Out of the total of 2453 patients, 281 needed care in the ICU (11.45%), and 270 stayed in the ICU for at least one day. These 270 patients with long stays in the ICU can be divided into 197 patients admitted from the hospital ward, and 73 admitted directly from the emergency service. On 7 May 2020, 57 of the long-stay ICU patients had died, 119 had been discharged to the hospital ward, and 43 were still in the ICU. **Figure S1** includes a flowchart related to the database. For the distribution of cases for different ages and sex, see **Table S1**.

**S2. Details on NP-MCM, KM and E estimates of lengths of stay until ICU admission from the hospital ward, death or discharge**

The estimator of S(t) in (eq2) is used to estimate the probability, p, of the event and the distribution of the times-to-event S_0_(t) using the relationships in (eq1) for the following lengths of stay.

*S2.1 Time in hospital ward (HW) until admission to ICU*

The goal is to estimate the probability that a patient in HW will need admission to ICU, and the distribution of the LoS in HW of those patients. The observations are {(t_i_, d_i_, x_i_), i = 1,…,n} with t_i_ the observed LoS in HW of all the patients, d_i_ indicates if the patient was admitted to ICU, and x_i_ the indicator of whether the admission to ICU was not observed because it will never happen since the patient died in HW or was discharged.

There were 2453 COVID-19 patients admitted to the hospital. In order to study the time in HW until ICU, we worked with the n = 2380 patients who were admitted first to HW, discarding the 73 patients who went to ICU directly from the emergency service. In the group of n = 2380 patients in HW, 197 of them required ICU. This gives an estimated empirical (E) probability of need for ICU p_emp_ = 197/2380 = 0.0828. But note that some of the patients still in HW at the end of the study would be admitted to ICU eventually, so the real probability is expected to be larger. The NP-MCM approach estimates that probability to be p_NP-MCM_ = 0.0845. The classical KM estimator considers n = 2380 patients in HW where 197 patients with admission to ICU is observed. This classical KM assumes that all the patients who had been admitted to HW will experience the event (admission to ICU) if followed for long enough, overestimating the LoS. This bias is partially corrected by the improved KM estimator, which takes into account that 1638 patients were discharged without ICU, and 328 died before being admitted to ICU. So it considers only n = 2380 – 1638 – 328 = 414 patients in HW with 197 patients where the event (admission to ICU) is observed. It still biases towards larger LoS, as patients still in HW by the end of the study are assumed to require ICU sometime in the future. The empirical estimator considers only n = 197 patients who were admitted to ICU, disregarding the information from the other right censored patients.

*S2.2 Time in hospital ward (HW) until death in HW*

The aim is to estimate the probability that a patient will die in HW, and the distribution of the LoS in HW of those patients. The observations are {(t_i_, d_i_, x_i_), i = 1,…,n} with t_i_ the observed LoS in HW of all the patients, d_i_ indicates if the patient died in HW, and x_i_ the indicator of whether the patient will not die in HW since he/she was discharged alive.

There were 2453 COVID-19 patients admitted to the hospital (into a hospital ward or the ICU). To study the time in HW until death, we worked with the n = 2183 patients who never required admission to ICU. In that group, 328 patients died, which gives an estimated empirical probability of death p_emp_ = 328/2183 = 0.1503. However, some of the 2183 patients were still in HW at the end of the study, and they might die eventually, so the probability of death in HW is expected to be larger. The NP-MCM approach estimates that probability to be p_NP-MCM_ = 0.1561.

Note that 1638 patients will never die in HW because they have been discharged; they are the known “cures” from death in HW. The classical KM estimator considers n = 2183 patients in HW with 328 observed events. The improved KM estimator takes into account that 1638 patients were discharged alive. Thus, it considers only n = 2183 – 1638 = 545 patients in HW with 328 patients where the event (death) is observed. The empirical estimator considers only the n = 328 patients whose event is observed, that is, those who died in HW, disregarding the information from the other patients.

*S2.3 Time in hospital ward (HW) until discharged without ICU*

The goal is to estimate the probability that a patient in HW will be discharged without requiring ICU, and the distribution of the LoS in HW of those patients. The observations are {(t_i_, d_i_, x_i_), i = 1,…,n} with t_i_ the observed LoS in hospital ward of all the patients, d_i_ indicates if the patient was discharged without need for ICU, and x_i_ the indicator of whether discharge will not be observed because the patient died before that event happened.

To study the time in HW until discharge, we worked with the n = 2183 patients in HW who did not need intensive care. In that group, 1638 were discharged, so the empirical estimator of the probability of discharge from HW without need for ICU is p_emp_ = 1638/2183 = 0.7503. However, there were patients still in HW at the end of the study, and many of them are expected to be discharged without admission to ICU, so the true probability of discharge from HW without ICU should be larger than p_emp_ =0.7503. The NP-MCM estimator of that probability is p_NP-MCM_ = 0.7953.

Note that 328 of the 2183 patients in HW will never be discharged because they died. They are the known “cures” from discharge. The classical KM estimator considers the n = 2183 patients in HW with 1638 patients where the event (discharge) is observed. The improved KM estimator takes into account that 328 patients died and will never be discharged. So it considers only n = 2183 – 328 = 1855 patients in HW with 1638 patients discharged. The empirical estimator considers only the n = 1638 patients discharged from HW, disregarding the information from the other patients.

*S2.4 Time in ICU until death in ICU*

The objective is to estimate the probability for a patient in the ICU of dying, and the distribution of the LoS in ICU of those patients. The observations are {(t_i_, d_i_, x_i_), i = 1,…,n} with t_i_ the observed time in ICU of all the patients, d_i_ indicates if the patient died in ICU, and x_i_ the indicator of whether the patient was discharged alive from ICU.

There were n = 270 patients admitted to ICU, and 53 of them died in ICU, so the empirical probability of death in ICU is p_emp_ = 53/270 = 0.1963. But in this group of n = 270 patients there were 42 patients still in ICU at the end of the study (which represent 42/270 = 15.55% of uncertainty), and 52 in HW discharged from ICU who might need ICU again. Note that any patient within these two groups might die in ICU eventually, so the number of deaths in ICU for these 270 patients is expected to be larger than the 53 observed deaths. As a consequence, the true probability of death in ICU should be larger than p_emp_ = 53/270 = 0.1963. The NP-MCM estimation of this probability is p_NP-MCM_ = 0.2222. These event percentages are corrected by the NP-MCM estimator, reducing the error down to 9.58%. Of course, the larger number of inpatients, the more efficient reduction of the error if the NP-MCM methodology is used for the estimation of the probabilities when there are patients with unobserved outcome.

To study the time in ICU until death, the NP-MCM estimator takes into account that some of the n = 270 patients in ICU will never die in ICU because they have been discharged from hospital (119) or died in HW after leaving ICU (4); they are the known “cures” from death in ICU. The classical KM estimator considers the n = 270 patients in ICU with n = 53 patients where the event (death) is observed. The improved KM estimator considers only n = 270 – 4 - 119 = 147 patients in ICU with 53 observed deaths. Finally, the empirical estimator considers only the n = 53 patients who died in ICU, disregarding the information from the other right censored times.

*S2.5 Time in ICU until discharged from ICU to HW*

The goal is to estimate the probability that a patient in ICU will be sent back to the hospital ward, and the distribution of the times in ICU of those patients. The observations are {(t_i_, d_i_, x_i_), i = 1,…,n} with t_i_ the time in ICU of all the patients, d_i_ indicates if the patient was discharged from ICU, and x_i_ the indicator of whether the patient died in ICU.

There were n = 270 patients who required ICU. To estimate the probability of discharge from ICU, the empirical estimator considers the 175 patients who were discharged from ICU (4 dead in HW after ICU, 52 still in HW and 119 discharged at home at the end of the study). This yields an estimated probability of discharge from ICU of p_emp_ = 175/270 = 0.6481. But some of the 42 patients still in ICU at the end of the study (53 patients died in ICU) may be discharged, so the real probability of discharge from ICU is expected to be slightly larger than 0.6481. The NP-MCM approach estimates that probability to be p_NP-MCM_ = 0.6820.

A total of 53 patients in ICU will never be discharged from ICU because they died in ICU; they are the known “cures” from discharge. The classical KM estimator considers the n = 270 patients in ICU with 175 observed events (discharge from ICU). The improved KM estimator takes into account that 53 patients died in ICU so they will never be discharged from ICU. It considers only n = 270 - 53 = 217 patients in ICU with 175 observed discharges. Finally, the empirical estimator considers only the n = 175 patients who have been discharged from ICU, disregarding the information from the other right censored patients.

**Table S2** shows the NP-MCM estimations of the time of hospitalization (which considers time in HW plus time in ICU) and the time in ICU for male and female patients, considering ages 40 and 70 years.

**S3. Density estimations for the different LoS.**

The density functions in **Figures S2 – S4** correspond to the survival estimates in **Figures 1, 2** and **S5** respectively. Both the NP-MCM and empirical estimators yield proper survival functions (they go down to zero as time *t* increases), so the corresponding density functions are proper (area equal to 1). Note that, however, when there are individuals who will not experience the final outcome, the KM is wrongly specified and the curves reach a plateau at the largest observed time, *t^1^_n_*. This has two important implications in the density function corresponding to the KM curve: (a) the area is not 1 but lower (the difference between 1 and the height of the plateau), so it should not be used for comparisons to proper density functions as those corresponding to the NP-MCM and E estimators; and (b) the KM curves estimate a large percentage of individuals (% = the value of the plateau) experiencing the event after the largest observed time, *t^1^_n_*, but the density function after *t^1^_n_* is zero. This zero value of the density function should not be interpreted as no events after that time *t^1^_n_* but simply lack of knowledge.

**S4. Model for simulating outbreak**

We considered a simulated outbreak with N = 1000 infected individuals. We assumed that the infection times, I_i_, i = 1, …, N, followed a log-Normal distribution, *Log-N(μ, σ),* with *μ* = 3.3 days and *σ* = 0.5 days [23]. For every i = 1, …, N, we simulated the sex *G_i_* (0 = male, 1 = female) and the age *A_i_* (years) of the *i-th* infected individual using the real distributions of the reported COVID-19 cases in Galicia on 7 May 2020 (for details in case counts see **Table S1**).

We defined H ᑕ {1, …, N} as the set of indices corresponding to infected subjects admitted in hospital. The trajectory of every patient *i* ∈ H was obtained by simulating the transitions between states of the state space *S* = {HW, ICU, D, Dis}, where *D* (death) and *Dis* (discharge) are terminal states, using the NP-MCM estimates of the probabilities p_j,k_ for j,k ∈ S. The duration times in states in *S* until transition to another state in *S* were also simulated using the Weibull distributions that best fitted the NP-MCM survival estimates.

The proposed model was used to perform a Monte Carlo simulation as follows. For every patient *i* = 1, …, N with age *A_i_* and sex *G_i_* the probability of admission in hospital *π(A_i_, G_i_)* is estimated from the reported and hospitalized cases in **Table S1**. Based on a U(0,1) random variate, U_i_, patient *i* is included in the set *H* of patients to be admitted into the hospital if U_i_ ≤ *π(A_i_, G_i_)*. This gave us the set *H*. The time since infection until hospital admission T_i_, of a patient *i* ∈ H was simulated from a normal distribution N(μ_i_, σ_i_) with μ_i_ = 12 - 0.05*A_i_* days and σ_i_ = 1.

When a patient is admitted into the hospital, the probability of going directly to ICU is p_H,ICU_ = 0.03, while the probability of staying in the hospital ward first is p_H,HW_ = 0.97. In the simulated model conditional on the age and sex of the patient, of those admitted in hospital ward, the probability of death without going to ICU is p_HW,D_(i) = 0.005exp(0.045*Ai*) and the time (days) to death follows a Weibull age-dependent and sex-dependent distribution, W(α_HW,D_(i), λ_HW,D_(i)), with parameters α_HW,D_(i) = 1.4 - 0.2*G_i_* and 1/λ_HW,D_(i) = 20exp(-0.008*A_i_*) for every *i* ∈ H. The probability that a patient admitted in hospital ward finally has to enter ICU is p_HW,ICU_ = 0.085, with the time (days) since hospital ward admission to ICU admission generated from a Weibull distribution W(α_HW,ICU_(i), λ_HW,ICU_(i)) with α_HW,ICU_(i) = 2.75 - 0.025*A_i_* and 1/ λ_HW,ICU_(i) = 2.5exp(0.02*A_i_*). As a consequence, the probability that a patient who was admitted to the hospital ward becomes discharged without entering ICU is p_HW,Dis_(i) = 0.915 - 0.005exp(0.045*A_i_*). The time (days) since hospital ward admission to discharge follows a Weibull distribution W(α_HW,Dis_(i), λ_HW,Dis_(i)), with α_HW,Dis_(i) = 1.75 (75 + 0.5*A_i_* - 11*G_i_*) / 100 and 1/ λ_HW,Dis_(i)=13 ( - 2.5 + 1.5*A_i_* – 7.5*G_i_* ) / 100. After being admitted in ICU a patient may die, with probability p_ICU,D_(i) = 0.0067exp((0.045 - 0.01*G_i_*)*A_i_*) or be transferred back to hospital ward, with probability p_ICU,HW_(i) = 1 - p_ICU,D_(i). Time from admission into ICU to death follows a Weibull distribution W(α_ICU,D_(i), λ_ICU,D_(i)), with parameters α_ICU,D_(i) = 0.8exp(0.009*A_i_*) and 1/ λ_ICU,D_(i) = 30exp(-0.012*A_i_*) for every *i* ∈ H. The distribution of the time since admission into ICU until return to ward is again Weibull W(α_ICU,HW_(i), λ_ICU,HW_(i)), with α_ICU,HW_(i) = 1.6 (1 + *G_i_*)exp(-0.003*A_i_*(1+4*G_i_*)) and 1/ λ_ICU,HW_(i) = 20exp(-0.003*A_i_*(1 -*G_i_*) - 0.22*G_i_*) for every *i* ∈ H. A summary of the considered Weibull parameters is presented in **Table S3** (see also **Figures S4** and **S5**). Note that for the different Weibull distributions, the shape parameters α(i) are truncated to be higher than 0.5, whereas the scale parameters 1/λ(i) are truncated to be higher than 1. All the estimated probabilities p_j,k_ for j,k ∈ S are truncated to fall between 0.05 and 0.95.

The effect of ignoring the dependence on age and sex can be shown by simulating an alternative model where all the probabilities and time-to-event distributions do not depend on these variables. More specifically, the probabilities of death, discharge and admission to ICU in a hospital ward are p_HW,D_ = 0.15, p_HW,Disc_ = 0.795 and p_HW,ICU_ = 0.085 respectively. The probabilities of death in ICU and discharge from ICU are p_ICU,D_ = 0.24 and p_ICU,HW_ = 0.76. The shape and scale parameters of the Weibull distributions, which no longer depend on age nor sex, are specified in **Table S3**.

**Figures and Tables**

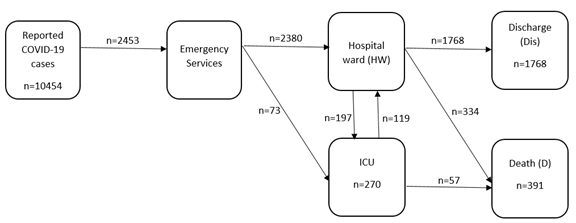

**Figure S1.** Flowchart of the confirmed COVID-19 cases reported in Galicia (Spain) from 6 March to 7 May 2020.

**Table S1.** Distribution of the total number of reported COVID-19 cases, and the number of reported cases hospitalized in Galicia (Spain) from March 6^th^ to May 7^th^, 2020.

|  | **Reported** | | **Hospitalized** | |
| --- | --- | --- | --- | --- |
| **Age** | **Women** | **Men** | **Women** | **Men** |
| +90 | 397 | 146 | 127 | 71 |
| 80-89 | 743 | 495 | 307 | 286 |
| 70-79 | 736 | 758 | 258 | 401 |
| 60-69 | 919 | 735 | 190 | 274 |
| 50-59 | 1150 | 705 | 118 | 161 |
| 40-49 | 1096 | 609 | 96 | 84 |
| 30-39 | 724 | 378 | 45 | 26 |
| 20-29 | 402 | 205 | 17 | 9 |
| 10-19 | 81 | 83 | 4 | 4 |
| 0-9 | 34 | 58 | 2 | 2 |
| Total | 6282 | 4172 | 1164 | 1320 |

**Table S2.** NP-MCM estimation of the mean, median and IQR of the duration of hospitalization (which considers time in HW plus time in ICU) and time in ICU, computed with the COVID-19 cases hospitalized in Galicia (Spain) from 6 March to 7 May 2020.

|  | **Duration of hospitalization** | | | **Time in ICU** | | |
| --- | --- | --- | --- | --- | --- | --- |
|  | **Mean^1^** | **Median** | **IQR** | **Mean^1^** | **Median** | **IQR** |
| Total | 16.80 | 11 | 7 - 19 | 23.94 | 17 | 9 - 38 |
| Women | 13.92 | 11 | 7 - 20 | 21.76 | 17 | 8 - 38 |
| Men | 18.41 | 10 | 6 - 17 | 23.88 | 18 | 10 - NA^2^ |
| 40 years | 15.36 | 9 | 6 - 16 | 21.82 | 16 | 8 - 28 |
| 70 years | 19.71 | 12 | 8 - 20 | 30.14 | 30 | 9 - NA^2^ |
| Women 40y | 8.47 | 8 | 5 - 10 | 16.56 | 15 | 8 - 19 |
| Men 40y | 16.84 | 10 | 6 - 18 | 23.58 | 16 | 8 - NA^2^ |
| Women 70y | 16.63 | 11 | 7 - 17 | 28.12 | 27 | 10 - NA^2^ |
| Men 70y | 19.60 | 12 | 7 - 21 | 24.28 | 17 | 8 - 38 |

^1^ Underestimate	^2^ NA = Not Available

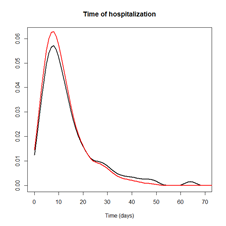

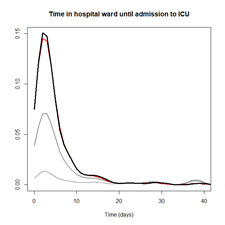

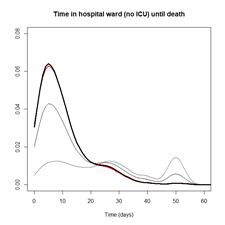

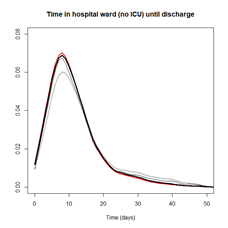

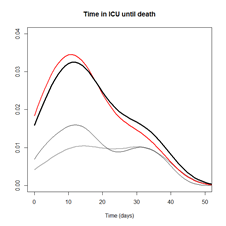

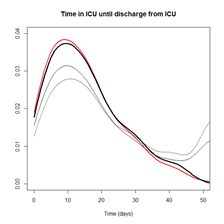

**Figure S2.** Estimates of the density function of LoS using NP-MCM (thick black line), KM with the complete dataset (thin grey line), KM with the reduced dataset (thin black line) and the empirical E estimator (red line) for all the COVID-19 hospitalized cases (n = 2453) in Galicia (Spain), when the LoS is the time of hospitalization (top left), time in hospital ward until admission to ICU (top right), time in hospital ward until death in hospital ward (middle left), time in hospital ward until discharge (middle right), time in ICU until death in ICU (bottom left) and time in ICU until discharge from ICU (bottom right).

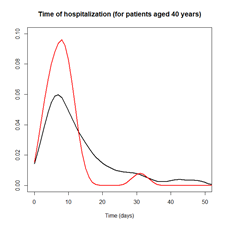

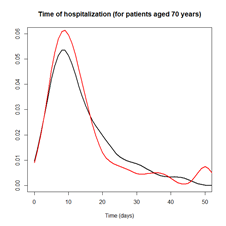

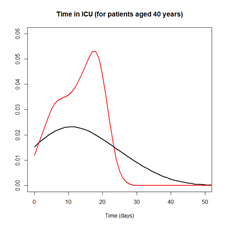

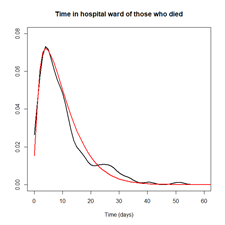

**Figure S3.** Estimates of the density function of the times of hospitalization which considers HW plus ICU (top) and time in ICU (bottom), incorporating the effect of the sex (male = black line, female = red line) and the ages 40y (left) and 70y (right) for all the COVID-19 hospitalized cases (n = 2453) in Galicia (Spain).

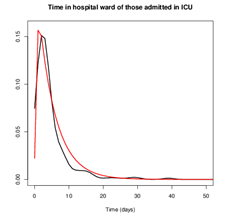

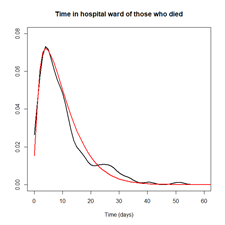

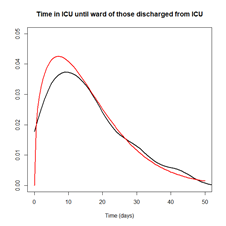

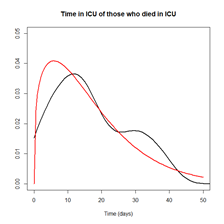

**Figure S4.** Estimation of the density function of different times-to-event using the NP-MCM (black) and Weibull distribution (red) unconditionally without taking into account sex and age of the patients, of the LoS in hospital ward until admission in ICU (top left), LoS in hospital ward until death (top right), LoS in hospital ward until discharge (middle left), LoS in ICU until death (middle right) and LoS in ICU until discharge to hospital ward (bottom left) for all the COVID-19 hospitalized cases (n = 2453) in Galicia (Spain).

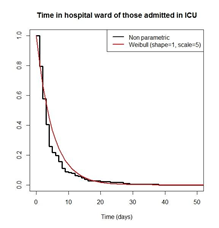

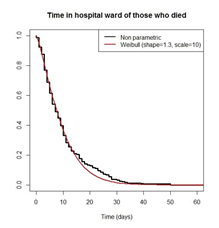

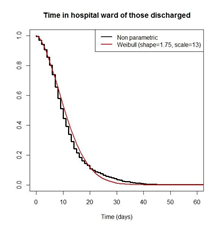

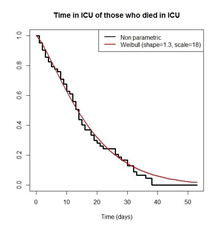

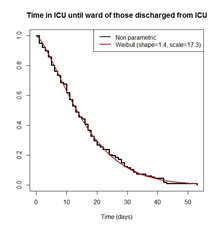

**Figure S5** Estimation, unconditionally without taking into account the age and sex of the patients, of the LoS in hospital ward until admission into ICU (top left), LoS in hospital ward until death (top right), LoS in hospital ward until discharge (middle left), LoS in ICU until death (middle right) and LoS in ICU until discharge to hospital ward (bottom left) using the NP-MCM (black) and Weibull distribution (red) for all the COVID-19 hospitalized cases (n = 2453) in Galicia (Spain).

**Table S3**. Parameters of the Weibull distribution fitted to the different times-to-event, based on the hospitalized COVID-19 patients in Galicia (Spain) from 6 March to 7 May 2020.

|  | **Model age (*A_i_*) and sex (*G_i_*) dependent** | | **Unconditional** | |
| --- | --- | --- | --- | --- |
| **Times** | α**(i)** | **1/**λ**(i)** | α | **λ** |
| HW to ICU | 2.75 - 0.025A_i_ | 2.5 exp(0.02A_i_) | 1 | 5 |
| HW to death | 1.4 - 0.2G_i_ | 20 exp(-0.008A_i_) | 1.3 | 10 |
| HW to discharge | 1.75(75 + 0.5A_i_ - 11G_i_)/100 | 13(-2.5 + 1.5A_i_ - 7.5G_i_)/100 | 1.75 | 13 |
| ICU to death | 0.8 exp(0.009A_i_) | 30 exp(-0.012A_i_) | 1.3 | 18 |
| ICU to HW | 1.6(1 + Gi) exp(-0.003A_i_(1 + 4G_i_)) | 20 exp(-0.003A_i_(1 – G_i_)-0.22G_i_) | 1.4 | 17.3 |

HW: Hospital ward; ICU: Intensive care unit

**R script for reproducing the results**

```
# This script contains the code for estimating:

# 1. The final outcome may happen for all the individuals
#    Example: Duration of hospitalization, time in ICU, etc
#    S(t) : survival function (Kaplan and Meier, 1958)
#    S(t|x): survival function conditioned on x (Beran, 1981)

# 2. The outcome is not experienced for a subgroup of individuals,
#    some of them clearly identified as being event-free, cure partially known
#    Example: Length of stay in hospital ward until admission in ICU/discharged alive/death
#    p: 	probability of experiencing the event (Safari et al, 2020)
#    S0(t): survival function of the individuals experiencing the event (Safari et al, 2020)


#######################################################################
# 1. THE FINAL OUTCOME MAY HAPPEN FOR ALL THE INDIVIDUALS
#---------------------------------------------------------------
#    S(t) : survival function (Kaplan and Meier, 1958)
#---------------------------------------------------------------

# Data frame - The observations are ordered based on the times Ti.
# time:   observed time to event
# status: indicator of whether the final outcome has been observed
data.real <- as.data.frame(cbind(time, status))

library(survival)
km_fit <- survfit(Surv(time, status) ~ 1, data = data.real)
km_fit$time   # Observed times
km_fit$surv   # Survival function S(t) evaluated at the observed times

#---------------------------------------------------------------
#    S(t|x): survival function conditioned on x (Beran, 1981)
#---------------------------------------------------------------
# Data frame - The observations are ordered based on the times Ti.
data.real <- as.data.frame(cbind(sex, age, time, status))
# sex:    sex of the individual
# age:    age of the individual
# time:   observed time to event
# status: indicator of whether the final outcome has been observed


# Covariate SEX: estimation of S(t|x) when x = 0 (men) and x = 1 (women).

library(survival)
km_fit.men <- survfit(Surv(time, status) ~ 1, data = data.real, subset = sex == 0)
km_fit.men$time   # Observed times
km_fit.men$surv   # Survival function S(t) evaluated at the observed
              	# times

km_fit.women <- survfit(Surv(time, status) ~ 1, data = data.real, subset = sex == 1)
km_fit.women$time   # Observed times
km_fit.women$surv   # Survival function S(t) evaluated at the
                	# times


# Covariate AGE: estimation of S(t|x) when x = 40 and x = 70.

library(npcure)
# Values of x = age where the survival function S(t|x) is estimated.
grid.age <- c(40, 70)
b_fit <- beran(x = age, t = time, d = status, dataset = data.real,
           	x0 = grid.age,
           	conflevel = 0.95,
           	cvbootpars = controlpars(hbound = c(0.2, 2), hl = 100))

# Survival function for age = 40 years:
S.40 <- b_fit$S$x40
# 95% confidence band
S.40.lower <- b_fit$conf$x40$lower
S.40.upper <- b_fit$conf$x40$upper

# Survival function for age = 70 years:
S.70 <- b_fit$S$x70
# 95% confidence band
S.70.lower <- b_fit$conf$x70$lower
S.70.upper <- b_fit$conf$x70$upper


# Covariates AGE and SEX: estimation of S(t|x) when x = 40 and male

grid.age <- 40
data_men <- subset(data.real, sex == 1)
b_fit_men <- beran(x = age, t = time, d = status, dataset = data_men,
            	x0 = grid.age,
            	conflevel = 0.95,
            	cvbootpars = controlpars(hbound = c(0.2, 2), hl = 100))
# Survival function for age = 40 years and sex = male:
S.men.40 <- b_fit_men$S$x40
# 95% confidence band
S.men.40.lower <- b_fit_men$conf$x40$lower
S.men.40.upper <- b_fit_men$conf$x40$upper

 #######################################################################
# 2. THE OUTCOME IS NOT EXPERIENCED FOR A GROUP OF INDIVIDUALS
# The survival function is S(t) = (1 - p) + p S0(t)
# p:     probability of experiencing the event
# S0(t): survival function of the individuals experiencing the event

# Data frame - The observations are ordered based on the times Ti:
# time:   observed time to event
# status: indicator of whether the final outcome has been observed
# cure:   indicator of whether the individual is known not to experience the event (cured)
data.real <- as.data.frame(cbind(time, status, cure))
#======================================================================
# Function that computes the nonparametric (NP) survival estimator
# when cure is partially known (CPK)
#======================================================================S_NPCPK <- function (data=data) {

  N <- nrow(data)

  # The observations are ordered based on the times Ti
  data.ot <- data[order(data[, 1]), ]
  t  <- data.ot[,1]
  d  <- data.ot[,2]
  nu <- data.ot[,3]
  cum.nu <- cumsum(nu)  # Number of known cures up to time Ti

  S <- rep(1, N)

  for (i in 2:N) {
    if(d[i]==0) {S[i] <- S[i-1]}
    if(d[i]==1) {S[i] <- S[i-1] * (1 - 1/(N - i + 1 + cum.nu[i-1]))}}

  p <- 1 - min(S)

  return(list(S, p, t))
}

#======================================================================
# Survival estimation
# S[[1]]: survival function S(t) evaluated at the observed times
# S[[2]]: probability (1 - p) of not experiencing the event
# S[[3]]: observed times Ti

S <- S_NPCPK(data.real)

# p : probability of experiencing the final outcome
p <- 1 - S[[2]]

# S0(t): Survival function of the individuals experiencing the event
S0 <- (S[[1]] - (1 - p))/p
#======================================================================
```
